# Individualized Functional Deviation Mapping: Linking Heterogeneous Structural Atrophy to Convergent Network Disruption in Preclinical Alzheimer’s Disease

**DOI:** 10.64898/2026.05.11.26352893

**Authors:** Iñigo Tellaetxe-Elorriaga, Antonio Jiménez-Marín, Ibai Díez, Asier Erramuzpe, Jesús M. Cortés

## Abstract

The preclinical phase of Alzheimer’s disease (AD) is characterized by profound biological and structural heterogeneity, challenging our ability to map early pathology onto large-scale brain networks. To address this fundamental challenge, we introduce Functional Deviation Maps (*π*_*z*_), an individualized neuroimaging framework for mapping participant-specific functional architecture to their unique structural atrophy landscape. By fitting a normative model to the voxel-based morphometry of amyloid-negative individuals, we extract personalized “atrophy seeds” (W-scores ≤ −1.96) for amyloid-positive patients, subsequently obtaining their resting-state seed-based connectivity (SBC). By standardizing these participant-level SBC maps against a healthy reference distribution, we show that, despite the highly variable spatial origins of structural atrophy, individual functional deviations converge into a common “atrophy network”. Spatial enrichment analyses show that the functional disruption is not random, but preferentially is dominated by the Default Mode Network. Furthermore, by projecting these populational functional deviations onto high-order cognitive topographies, we find a considerable alignment with the brain’s fundamental unimodal-transmodal and external-internal attentional gradients. Overall, the *π*_*z*_ framework transcends conventional group-level averages, offering a highly personalized, biologically meaningful signature of system-level network vulnerability in the earliest stages of AD.

## 1. Introduction

While Alzheimer’s disease (AD) is now anchored by a strict biological definition, its clinical and brain structural abnormalities remain heterogeneous, characterized by striking inter-individual variability in temporal onset, phenotypic trajectories, and the spatial topography of neuropathology (Ferreira et al., 2020; Vogel et al., 2021). This heterogeneity is already evident in the preclinical phase of AD (Pelkmans et al., 2024; Xu et al., 2024), operationally defined within the AT(N) framework(Jack Jr. et al., 2024; Jack et al., 2016), where “A” refers to amyloid-*β* (A*β*) pathology (typically measured via PET imaging or CSF A*β*42/40 ratio), “T” to tau pathology (phosphorylated tau in CSF or tau PET), and “N” to neurodegeneration or neuronal injury (e.g., structural atrophy on MRI or hypometabolism on FDG-PET). Individuals classified as cognitively unimpaired (CU) but biomarker-positive for amyloid (A^+^), with variable tau (T^±^) and variable neurodegeneration (N^±^) represent a biologically defined stage in which molecular pathology is present in the absence of overt clinical symptoms (Sperling et al., 2023). Characterizing how this early biological heterogeneity maps onto network-level brain alterations is crucial both for optimizing preclinical trial enrichment and explaining divergent clinical trajectories (Hampton et al., 2020; Ossenkoppele et al., 2022).

At the structural level, gray matter atrophy exceeding age-expected loss is a hallmark of AD, yet its spatial patterns are highly heterogeneous in the preclinical phase (Lorenzini et al., 2025). At the functional level, the Default Mode Network has shown to be highly vulnerable (Cha et al., 2026; Guzmán-Vélez et al., 2022; Ingala et al., 2021; Zhukovsky et al., 2023) but AD-related functional neuroimaging often extends to other association networks and limbic systems with profiles that vary significantly across individuals (Grothe et al., 2016). Crucially, structural degeneration and functional connectivity alterations exhibit only partial spatial overlapping. This raises the fundamental question of whether the anatomically heterogeneous patterns of preclinical AD atrophy map onto a common underlying functional network architecture, potentially reflecting shared system-level vulnerability despite regional anatomical variability.

Prior work in symptomatic AD and related network-mapping studies suggests that regional gray-matter loss is shaped by the organization of large-scale functional networks (Ossenkoppele et al., 2015), so that spatially distinct atrophy regions often map onto a common connectivity pattern in healthy brains (Tetreault et al., 2020). This type of approach has been termed Atrophy Network Mapping, representing a natural extension of Lesion Network Mapping by substituting focal lesions with regions of atrophy (Fox, 2018; Jimenez-Marin et al., 2022). However, these approaches typically rely on normative connectivity data, estimating how atrophy regions connect in an average brain rather than quantifying how connectivity in the affected individual deviates from a healthy reference. This limitation is important because functional connectivity shows substantial inter-individual variability and stable subject-specific organization (Finn et al., 2015; Mueller et al., 2013).

Recent methodological work has further highlighted that when atrophy sites (or lesions) are highly spatial heterogeneous, the underlying common network connected to all lesions is strongly shaped by dominant properties of the reference connectome, and in particular, the map of degree, ie., when averaging across many distinct lesions, the resulting population-level disconnection maps converge toward the degree brain map of the connectome (Van Den Heuvel et al., 2026). However, by implementing a dual strategy of sensitivity and specificity, it is possible to demonstrate that beyond degree-driven effects, there exist normative disconnection patterns that correlate with specific symptoms independently of degree maps (Meng et al., 2026; Siddiqi et al., 2026). Together, these considerations motivate moving forward toward the need for individualized metrics that integrate participant-specific structural pathology with participant-specific functional architecture.

To address this gap, we introduce the Functional Deviation Map (*π*_*z*_) framework, an individualized approach that anchors resting-state functional connectivity to each participant’s atrophy-defined seed and quantifies deviations relative to a reference distribution derived from amyloid-negative cognitively unimpaired individuals. We show that subject-specific functional deviations from normality can be measured directly at the individual level and that those deviations converge across A^+^CU participants. Using data from the A4 and LEARN studies, we therefore tested whether gray-matter atrophy in A^+^CU individuals delineates a consistent atrophy-related functional network, whether that network shows systematic functional deviation relative to an A^−^T^−^CU reference, and whether such deviations relate to tau pathology and cognitive performance and organization.

## 2. Materials and Methods

### 2.1. Participants

Data corresponded to cognitively unimpaired (CU) older adults enrolled in the publicly accessible Anti-Amyloid Treatment in Asymptomatic Alzheimer’s Disease (A4; ClinicalTrials.gov NCT02008357) trial and its observational companion study, the Longitudinal Evaluation of Amyloid Risk and Neurodegeneration (LEARN; ClinicalTrials.gov NCT02488720), available through the LONI Image and Data Archive (https://ida.loni.usc.edu/login.jsp). The A4 trial is a double-blind, placebo-controlled secondary prevention study designed to determine whether the anti-amyloid monoclonal antibody solanezumab can slow cognitive decline in CU older adults with elevated cerebral amyloid-*β* (A*β*). LEARN follows individuals who did not meet the A*β* elevation criteria for A4 and were therefore excluded from randomization but remained in an observational cohort. Participants were required to be between 65 and 85 years old, live independently, and be cognitively unimpaired, defined by a Clinical Dementia Rating (CDR) global score of 0, a Mini-Mental State Examination (MMSE) score between 25 and 30, and a Wechsler Memory Scale Logical Memory Delayed Recall (LMDR) score of 6–18. Individuals were excluded if they had a diagnosis of cognitive impairment or dementia, were taking Alzheimer’s disease medications, had unstable medical illnesses, or had significant anxiety or depressive symptoms that might make disclosure of A*β* imaging results unsafe. All study procedures were approved by the Institutional Review Board at each site, and written informed consent was obtained from all participants before screening.

For our study, we defined the reference group as the CU amyloid negative and plasma pTau217 negative (A^−^T^−^CU) individuals, and the clinical group as the CU amyloid positive (A^+^T^±^CU), independent of plasma pTau217 positivity (T^+^) or negativity (T^−^). Amyloid and tau status were determined by determining in the amyloid PET values of SUVr < 1.10 for the A^−^T^−^CU reference group and SUVr > 1.15 for the A^+^T^±^CU clinical group, following(Sperling et al., 2023). Those participants in the intermediate range of 1.10 < SUVr < 1.15 were discarded to avoid false positive and false negative errors. To determine tau positivity, a cutoff of pTau217 < 0.2 U/ml was applied based on previous studies(Sperling et al., 2024). This cutoff corresponds to individuals who remained cognitively stable over 288 weeks, as assessed by longitudinal PACC scores. As a result, our sample included 245 A^−^T^−^CU participants and 544 A^+^T^±^CU participants. For simplicity, hereafter we refer to these groups as A^−^and A^+^, respectively. The A^+^group comprised both T^+^ (N=141) and T^−^ (N=403) participants. The demographics of the analyzed group are shown in Table 1.

**Table 1:**
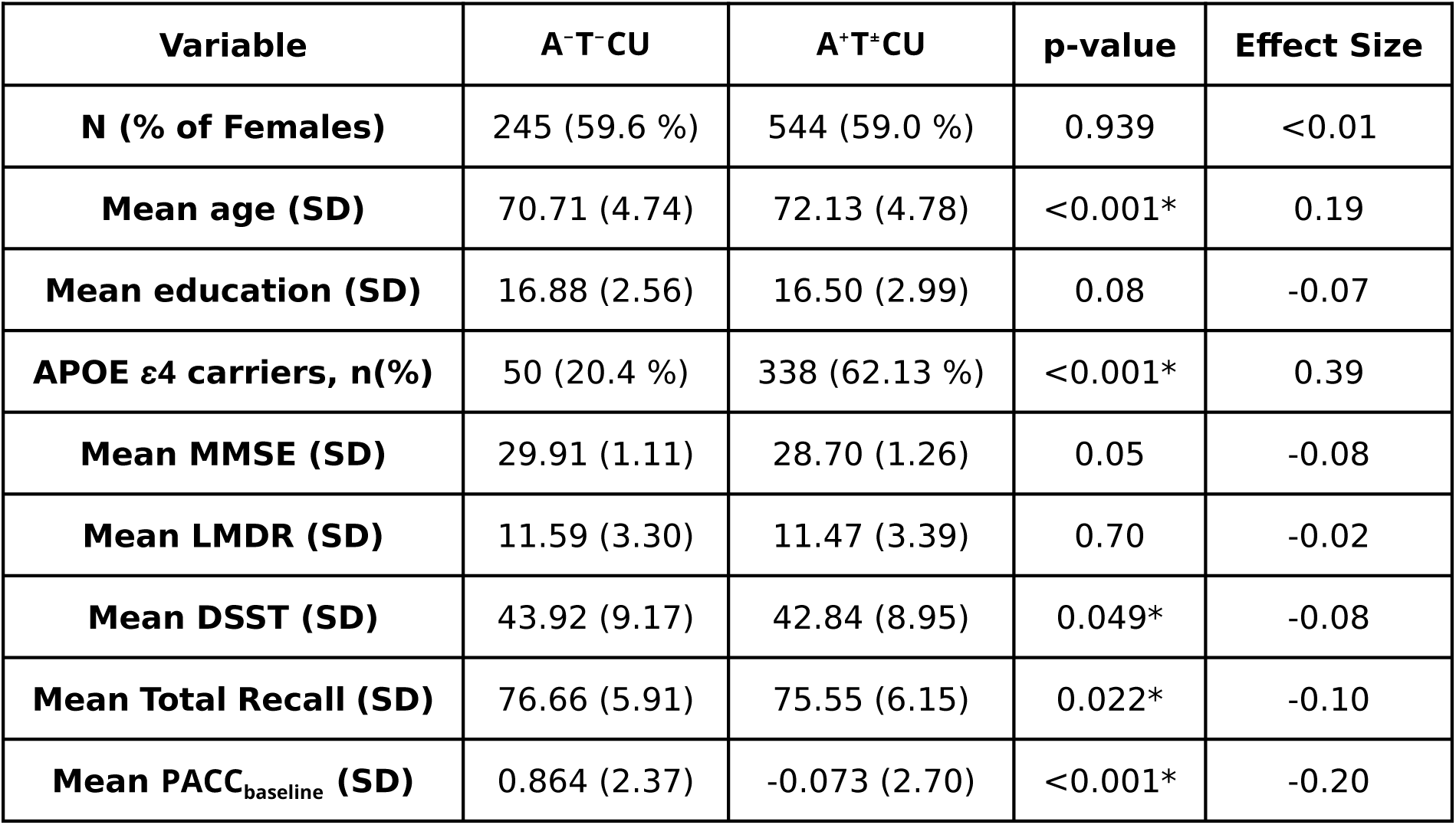
Participant demographics in the defined clinical groups and their cognitive performance. Abbreviations: APOE *ε*4 carriership, Mini-Mental State Examination (MMSE), Logical Memory Delayed Recall (LMDR), Digit Symbol Substitution Test (DSST). Statistically significant (∗). Effect sizes are given as rank-biserial correlation *r* and the *ϕ* coefficient for numerical and categorical variables, respectively.

### 2.2. Neuroimaging data and preprocessing

We included both T1-weighted structural and resting-state fMRI images acquired on 3T MRI scanners with standardized ADNI protocols(Jack Jr. et al., 2010) across all participating study sites. Briefly, The T1-weighted anatomical scans were collected using MPRAGE sequences with TR ranging from 2300–7636 ms, TI from 400–900 ms, and flip angles of 9–11°, providing high-resolution 1 mm isotropic structural images. The resting-state fMRI data consisted of approximately 6.5-minute echo-planar imaging (EPI) scans with TR values between 2925–3520 ms, TE of 30 ms, and flip angles of 80–90°, acquired on GE, Philips, and Siemens scanners.

To preprocess the neuroimaging data, we used the FMRIB Software Library v6.0.4 (FSL) (Smith et al., 2004) and MATLAB 2021b. The anatomical T1-weighted MRI preprocessing pipeline included: re-orientation to right-posterior-inferior (RPI); alignment to anterior and posterior commissures; skull stripping; gray matter, white matter and cerebrospinal fluid segmentation; and computation of a non-linear transformation between individual skull-stripped T1 and 2 mm resolution MNI152 template images.

The functional MRI preprocessing pipeline included: slice time correction; reorientation to RPI; re-aligning functional volumes with a rigid body transformation (6 parameters linear transformation); computation of the transformation between individual skull-stripped T1 and mean functional images; intensity normalization; transformation to MNI standard space and smoothing with an isotropic Gaussian kernel of 8-mm FWHM. Then ICA-AROMA was used, a data-driven method, to identify and remove motion-related independent components from fMRI data. Then removal of confounding factors from the data using linear regression including 6 motion-related covariates, linear and quadratic terms, and five components each from the lateral ventricles and white matter. Band-pass filtering (0.01–0.08 Hz) was applied to reduce low-frequency drift and high-frequency noise. Head motion was quantified using realignment parameters obtained during image preprocessing, which included 3 translation and 3 rotation estimates. Scrubbing of time points with excess head motion interpolated all time points with a frame displacement > 0.5 mm.

### 2.3. Neuroimaging analyses

#### 2.3.1. Voxel-Based Morphometry and atrophy seeds

To quantify GM volume (GMv), the FSL voxel-based morphometry (FSLVBM) pipeline was used: the individual subject GM partial volume estimates were registered to MNI152 standard space using non-linear registration. A left–right symmetric, study-specific GM template was subsequently created by averaging the registered images and flipping them along the x-axis (antero-posterior). Finally all native GM images were non-linearly registered to the study-specific template and modulated to correct for local expansion or contraction induced by the non-linear component of the spatial transformation. The modulated GM images were smoothed using an isotropic Gaussian kernel with a *σ* = 3.

For determining atrophy seeds, a voxel-wise general linear model (GLM) was fitted for the GM using the images only from the A^−^group using age, age squared, gender, and total intracranial volume as covariates. Next, we calculated a voxel-wise W-score for GM in each patient in the A^+^ group. A W-score, used before in(Ossenkoppele et al., 2015), is simply a Z-score adjusted for the mentioned covariates. W-score maps were then thresholded at W ≤ −1.96 as done in previous studies and binarized to obtain the “atrophy seeds” of each participant (Ossenkoppele et al., 2015). All analyses were performed in the MNI152 standard space. A graphical summary for the atrophy seed estimation is shown in Figure 1.

**Figure 1:**
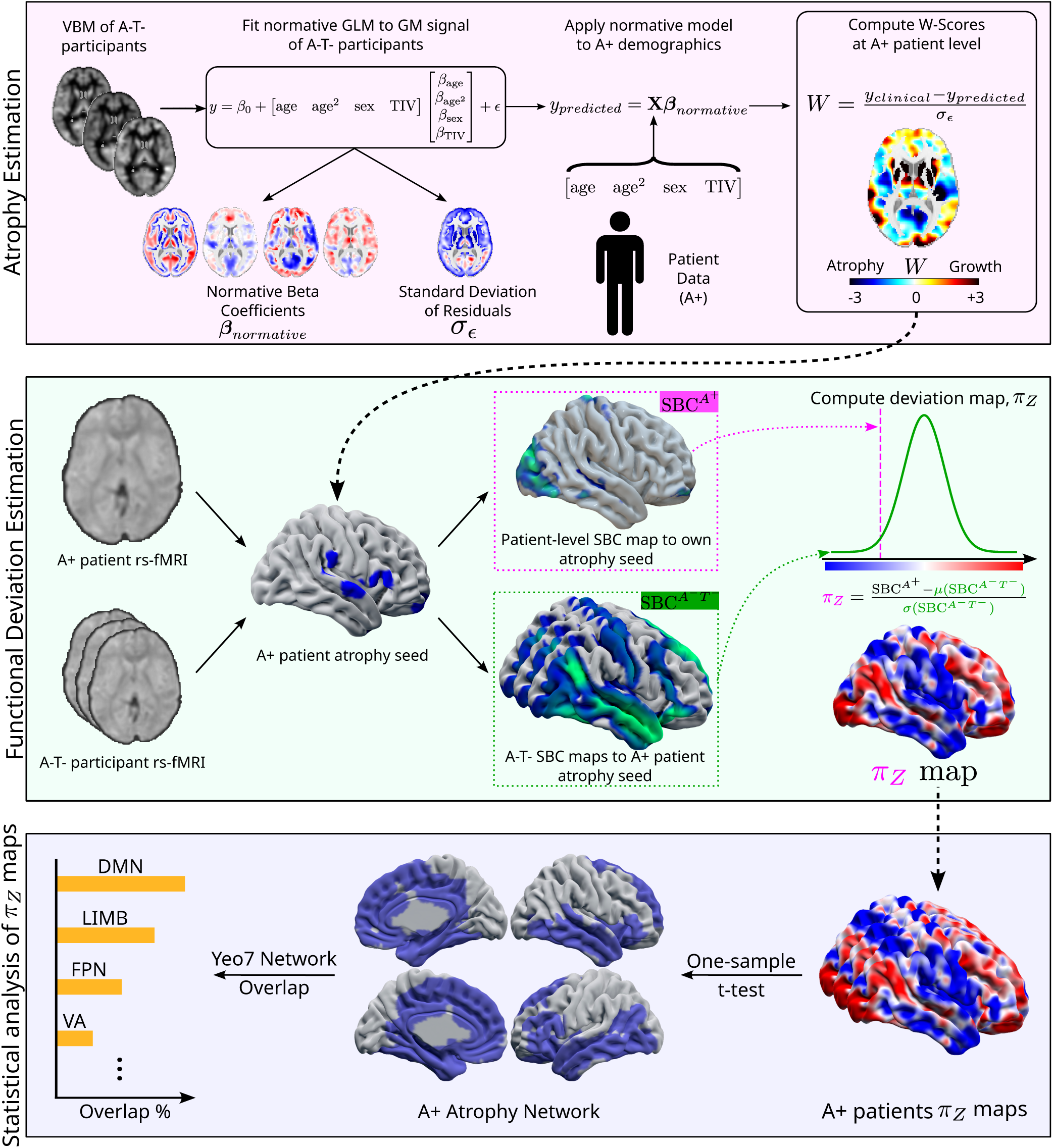
Deviation Maps Calculation Pipeline. Purple: Voxel based morphometry signals from the healthy reference (A^−^T^−^ CU) group are used to fit a voxelwise General Linear Model, correcting for age, age^2^, sex, and Total Intracranial Volume. With the fitted model, W-scores are computed as the difference between the VBM signal of the A^+^ participant and their predicted VBM signal by the model, divided by the standard deviation of the model residuals. W-score maps are thresholded at W ≤ −1.96 and binarized to obtain the atrophy seeds. Green: Seed Based Connectivity is computed with the atrophy seeds, using the A^+^ participant and the A^−^T^−^ group rs-fMRI images, producing a collection of SBC maps. Then, the 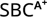 map is z-scored respect to the 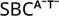 maps, obtaining the atrophy functional deviation map **π**_*z*._ Blue: With the **π*_*z*_* maps for all A^+^ participants, a voxelwise one-sample t test is run to check if the deviations are consistent across the whole A^+^ group. After correcting for multiple comparisons, statistically significant (*p* < 0.05) voxels are overlaid with the Yeo 7 network parcellation to get a functional description of the discovered networks.

#### 2.3.2. Functional deviation maps

To provide an individualized measure of atrophy driven network disruption, we quantified, for each A^+^participant, the extent to which their functional connectivity to their own atrophy seeds deviates from a reference distribution derived from A^−^individuals. To this end, we performed two seed-based connectivity (SBC) analyses. First, for each A^+^participant, we computed Pearson correlation coefficients (*ρ*) between the mean time series of the atrophy seed and the time series of every other voxel in the brain using their rs-fMRI data. This resulted in an 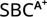 map. Then, using the same atrophy seed defined for the A^+^ participant, we applied the same procedure to each A^−^ participant, computing *ρ* maps from their rs-fMRI data. This resulted in one 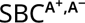 map per A^−^ participant and the seed defined in the A^+^ participant. All *ρ* maps were then Fisher-transformed using the inverse hyperbolic tangent function. Next, we computed the voxel-level mean (*µ*_A−_) and standard deviation (*σ*_A−_) across all 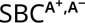 maps and used these to derive the voxel-level deviation map (*π*) for the A^+^ participant, as defined in Equation 1:

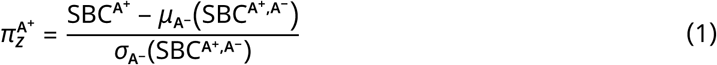

Figure 1 shows major steps for the computation of *π*_*z*_.

To investigate the existence of consistently functionally deviated regions, two voxel-wise one-sided one-sample t-tests were performed on all 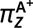 maps. To correct for multiple comparisons while accounting for the intrinsic spatial correlations found in statistical maps in all the fitted GLMs, we employed a 3D cluster correction method. Building a null distribution of randomly generated 3D clusters using AFNI’s `3dClustSim`, we estimated the smoothness of the residuals in the fitted GLM using AFNI’s `3dFWHMx`, running 5,000 thousand Monte Carlo simulations at increasing significance levels (*α* = [0.05, 0.04, 0.03, 0.02, 0.01, 0.005, 0.002, 0.001, 0.0005, 0.0002, 0.0001]). With this, we established a minimum cluster size for each significance level. Then, the Z-score maps corresponding to the one-sample t-tests were thresholded at the established *α* levels, and all the clusters exceeding the computed minimum cluster sizes were considered statistically significant.

For functional characterization of the population maps, to describe the regions passing statistical significance thresholds functionally, we computed their enrichment with the 7 resting state canonical networks described by (Thomas Yeo et al., 2011). For each canonical resting network *R*_*n*_, the number of overlapping voxels was normalized either by the size of the canonical network or by the size of the atrophy network. To account for potential inflation of overlap in larger networks, spatial enrichment (E_*n*_) was computed using the location quotient (see Equation 2). Intuitively, larger networks are penalized in this metric, as they have a higher probability of containing voxels from the mask (*M*), and vice versa for smaller networks. Values of E_*n*_ > 1 indicate that a network is enriched beyond chance expectation, whereas values close to or below 1 indicate no enrichment or depletion. We define the enrichment E_*n*_ between a given mask *M* and a cannonical resting state network *R*_*n*_ as:

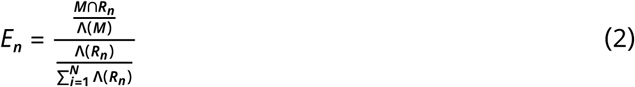

where 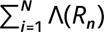 denotes the total volume of all canonical resting-state networks, and Λ represents the volume of a given mask or network. The numerator in Equation 2 corresponds to the probability that a voxel from the mask *M* falls within the resting-state network *R*_*n*_, whereas the denominator represents the baseline probability of observing *R*_*n*_ within the whole-brain space. Thus, the enrichment can be interpreted as the relative likelihood that *M* overlaps with *R*_*n*_ compared to chance, given the spatial extent of *R*_*n*_.

#### 2.3.3. Association between functional deviation maps and clinical variables

The clinical variables considered in this study included: PACC scores at specific timepoints (e.g., week 0, week 240), longitudinal changes in PACC (Δ_PACC_ = PACC_*t*=final_ − PACC_*t*=0_), baseline pTau217 levels (week 0), baseline Centiloid values (week 0), and the interaction between pTau217 and Centiloid at baseline. To assess the clinical associations of deviation maps, we fitted voxel-wise GLMs using *π*_*z*_ as the response variable, while adjusting for covariates including years of education, age, and sex.

#### 2.3.4. Correlation between population functional deviations and cognitive architecture

Finally, we asked whether the high order cognitive topography of the brain is related to the populational functional deviation, as measured by the Z statistic for testing 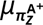 < 0. To investigate this question, we employed Neurosynth to obtain probability maps for a set of psychological terms (Yarkoni et al., 2011) that were limited to those shared between Neurosynth and the Cognitive Atlas (Poldrack et al., 2011). For maintaining comparability to other studies, we employed the same list as in (Shafiei et al., 2020). This resulted in a total of 123 terms. We then calculated the first two principal components of the 123 term association maps and computed their correlation with the populational map of functional deviation. To determine which spatial patterns of psychological terms most closely match the functional deviations linked to preclinical atrophy, the correlation between the top 10 concept maps with the highest loadings in their PC1 and PC2 decompositions was measured. To account for spatial autocorrelation, we parcellated the brain maps with 2,507 regions of interest as given by (Jimenez-Marin et al., 2023; 2024) and generated 5,000 null-maps for each PC1 and PC2 maps using NiMARE (Salo et al., 2023) (Figure 3, *α* = 0.05 Bonferroni corrected).

## 3. Results

### 3.1. Deviation Maps Pipeline

The full analysis pipeline is summarized in Figure 1, from the estimation of structural atrophy seeds to the calculation of functional deviation maps and their statistical analysis. In the first stage (top panel), VBM signals from the A^−^ reference group were used to fit a voxelwise normative GLM, including age, age squared, sex, and total intracranial volume as covariates. This model yielded voxel-wise normative coefficients (*β*_normative_) and residual variability (*σ*_*ε*_), which were subsequently used to compute W-score maps at the individual A^+^level. These W-scores, defined as the difference between observed and model-predicted gray matter signal normalized by *σ*_*ε*_, spanned a symmetric range (approximately −3 to +3), with negative values indicating atrophy. Thresholding at W-score ≤ −1.96 defined subject-specific atrophy seeds.

In the second stage (middle panel), these atrophy seeds were used to derive SBC maps. For each A^+^participant, an 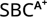 map was computed from their own rs-fMRI data, capturing the functional connectivity between the atrophy seed and the rest of the brain. In parallel, the same seed was applied to all A^−^ participants to generate a distribution of 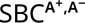 maps. These maps were Fisher-transformed and used to estimate a reference distribution (mean and standard deviation) of connectivity values. The functional deviation map *π*_*z*_ for each A^+^ participant was then computed as a voxel-wise Z-score, comparing 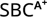 against the reference. As illustrated in Figure 1, *π*_*z*_ values reflect deviations along a continuous axis, where positive and negative values indicate increased or decreased connectivity relative to the normative distribution.

### 3.2. Group deviations preferentially target transmodal networks

At the group level (bottom panel), *π*_*z*_ maps from all A^+^ participants were entered into a voxel-wise one-sample t-test to identify regions showing consistent functional deviation across subjects. The resulting statistical maps revealed non-random spatial structure, with negative deviations converging across participants. Overlap analysis with the Yeo 7network parcellation further showed that these deviations were preferentially localized within canonical large-scale networks, including default mode (DMN), limbic (LIMB), frontoparietal (FPN), and visual/attention (VA) systems, as quantified by the percentage overlap shown in the bar plot.

The spatial distribution of consistent functional deviations across A^+^participants revealed a well-defined atrophy-related network encompassing both cortical and subcortical regions (Figure 2). These regions predominantly exhibited negative deviation values *π*_*z*_ < 0, indicating reduced functional connectivity relative to the A^−^ reference. The resulting pattern was not diffusely distributed but instead formed a coherent large-scale network, with prominent involvement of medial and lateral cortical areas as well as subcortical structures. Quantitative overlap with the Yeo 7-network parcellation showed a preferential involvement of higher-order association systems. The DMN showed the largest overlap, reaching 31.0% when referenced to the network (i.e., percentage of the DMN covered by the mask) and 30.22% when referenced to the mask (i.e., percentage of the mask falling within the DMN). Limbic regions also showed substantial overlap (20.47% to network; 7.65% to mask), followed by the Ventral Attention Network (12.12% to network; 5.36% to mask) and the Frontoparietal Network (11.11% to network; 7.02% to mask).

**Figure 2:**
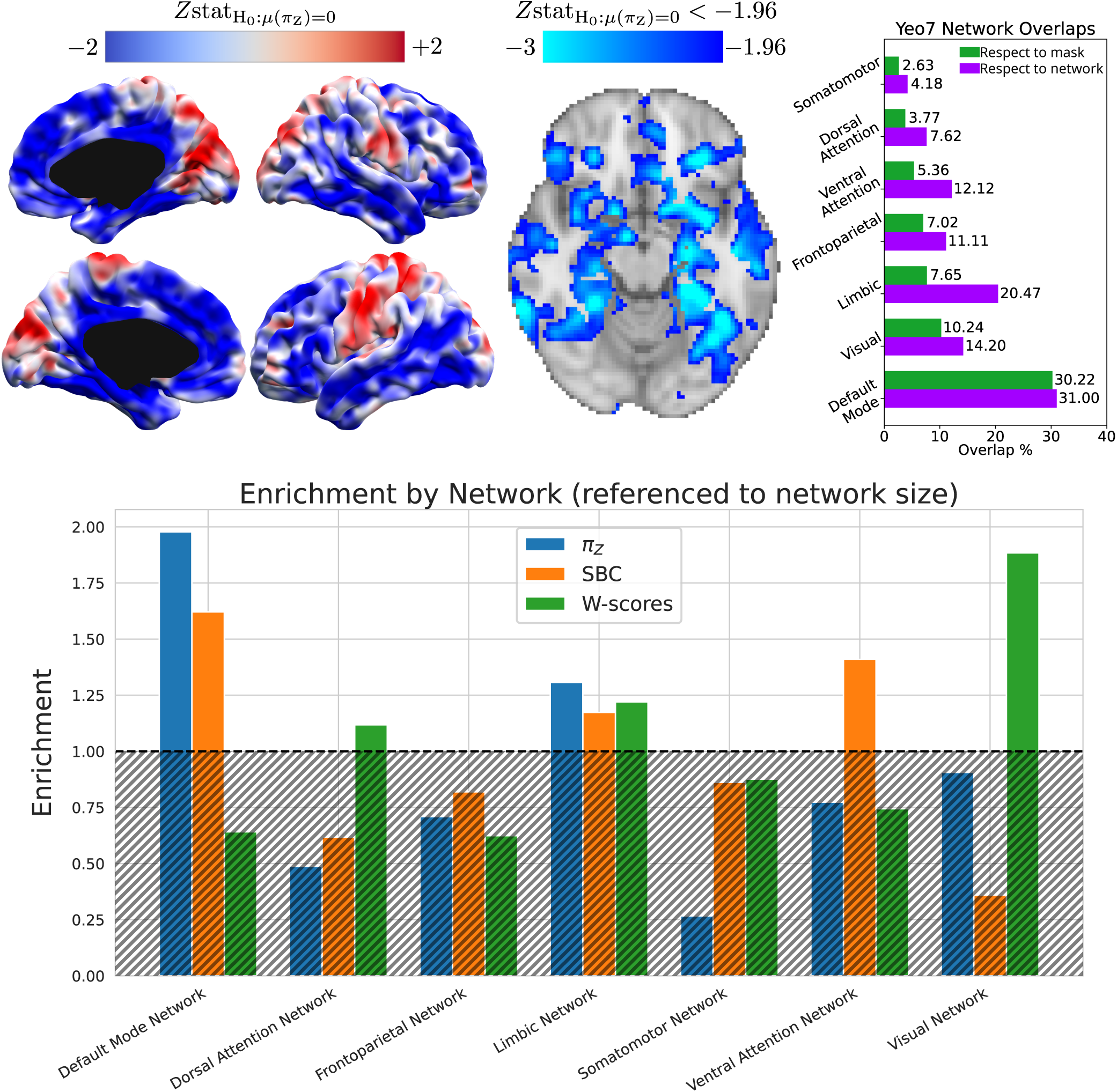
<u>Top left</u>: Cortical and subcortical populational deviation map of the discovered atrophy network <u>Top center</u>: Voxels with statistically significant negative deviation values across A+ participants in the populational deviation map. <u>Top right</u>: Differential overlap percentages with the canonical 7-network parcellation from (Thomas Yeo et al., 2011), referenced to the size of the atrophy network or to the size of the resting state network mask. <u>Bottom panel</u>: Enrichment (*E*_*n*_) values per Yeo 7 network of the statistically significant voxelwise one-sample t-tests performed in the W-score, SBC, and *π*_*z*_ maps. The dotted horizontal line at *E*_*n*_ = 1 represents the null expectation, and the region above it represents enriched network masks.

In contrast, primary systems exhibited lower involvement. For example, in the Somatomotor network, only 2.63% of the mask was located within this network (mask-referenced overlap), while 4.18% of the Somatomotor network was covered by the mask (network-referenced overlap). Similar patterns were observed for the Dorsal Attention network (3.77% of the mask; 7.62% of the network) and, to a lesser extent, the Visual network (10.24% of the mask; 14.20% of the network). These results indicate that the atrophy-related functional deviations preferentially target associative and transmodal systems rather than unimodal sensory-motor networks.

Network-level enrichment analysis is shown in Figure 2, bottom panel. Focusing on enrichment values above the null expectation (E_*n*_ > 1), *π*_*z*_ maps showed the strongest enrichment in the DMN (E_*n*_ ≈ 2.0) and, to a lesser extent, in the Limbic network (E_*n*_ ≈ 1.3), indicating that functional deviations relative to the normative reference preferentially involve these systems. In parallel, SBC maps (reflecting within-group atrophy connectivity patterns in A^+^participants) also showed enrichment in the DMN (E_*n*_ ≈ 1.6) and Ventral Attention network (E_*n*_ ≈ 1.4), consistent with a distributed large-scale functional reorganization at the group level. In contrast, W-score maps, capturing purely morphometric (atrophy-driven) effects, showed their strongest enrichment in the Visual network (E_*n*_ ≈ 1.9), with comparatively weaker contributions in higher-order associative systems.

Taken together, these results indicate a dissociation between structural and functional organization. While atrophy (W-scores) is primarily concentrated in posterior sensory regions such as the Visual network, the dominant large-scale pattern across the brain is better captured by functional measures, namely SBC (within-group connectivity) and *π*_*z*_ (functional deviation from the normative reference). In particular, *π*_*z*_ isolates a networklevel signature that emphasizes higher-order associative systems, especially the DMN and Limbic networks, suggesting that the impact of atrophy extends beyond its anatomical locus through distributed functional alterations.

### 3.3. Group deviations align with the brain’s cognitive topography

To further characterize the large-scale organization of functional deviations, we examined their alignment with canonical cognitive gradients (Figure 3). The first principal component (CogPC1, *λ*_explained_ = 15.02%), reflecting the unimodal–transmodal axis, showed a significant positive correlation with the A^+^ deviation maps (*ρ* = 0.33, p = 0.005), confirming that functional deviations are systematically organized along the unimodal–transmodal hierarchy, with greater involvement of transmodal regions. At the level of cognitive concepts, significant positive correlations were observed for higher-order affective processes, including valence, emotion, and fear (all p < 0.05), indicating their alignment with the observed deviation pattern. In contrast, the concept of action showed a significant negative correlation (p < 0.05), consistent with reduced involvement of sensorimotor domains.

**Figure 3:**
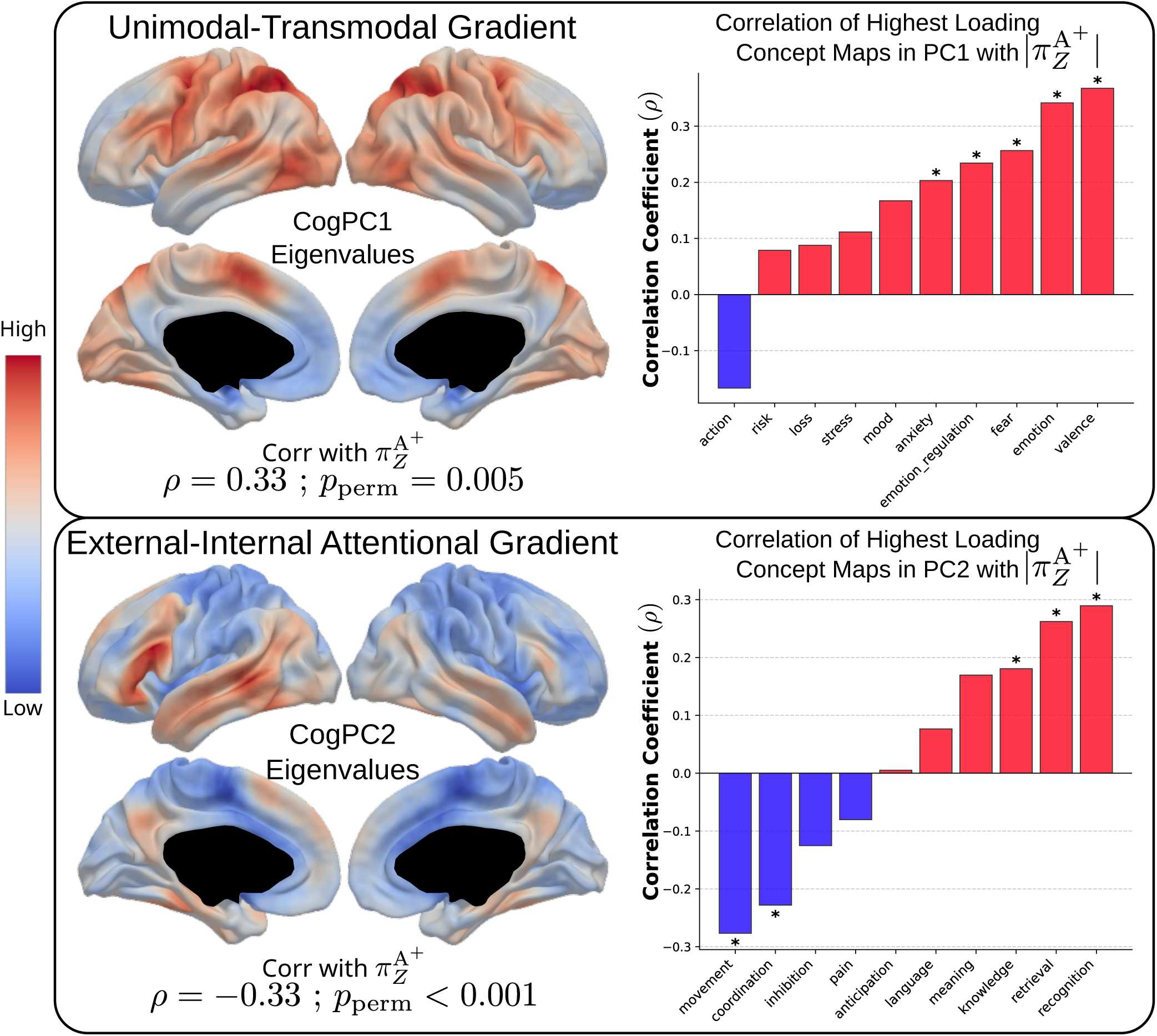
<u>Left</u>: CogPC1 and CogPC2 maps derived as described in the `neuromaps` Python package (Markello et al., 2022) and their correlation with A^+^ population 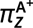 maps. <u>Right</u>: correlations between the population |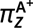| map and the top 10 cognitive concepts with the highest weights in the CogPC1 and CogPC2 principal component decompositions. Asterisks (∗) indicate **p**_perm_ < 0.05.

The second principal component (CogPC2, *λ*_explained_ = 9.30%), capturing the external–internal attentional axis, showed a significant negative association with *π*_*z*_ maps (*ρ* = −0.33, p < 0.001), indicating that functional deviations preferentially align with internally oriented processing systems. Consistently, significant positive correlations were observed for internal cognitive processes such as recognition, retrieval, knowledge, and meaning (all p < 0.05). Conversely, motor and control-related concepts—including movement, coordination, and inhibition—showed significant negative correlations (all p < 0.05). Together, these results demonstrate that functional deviations in A^+^ participants are selectively aligned with transmodal and internally oriented cognitive systems, while sensorimotor processes are significantly underrepresented.

## 4. Discussion

In this study, we present a personalized neuroimaging framework to characterize functional vulnerability at the individual level in preclinical AD. Here, personalized specifically refers to the ability of the framework to operate at the level of a single individual rather than relying on group-averaged representations. By introducing Functional Deviation Maps (*π*_*z*_), we move beyond conventional group-level analyses to quantify how subject-specific atrophy seeds reshape functional connectivity relative to a normative A^−^reference. This approach enables a direct assessment of how early structural alterations propagate into individualized patterns of functional disruption.

This framework extends prior atrophy network mapping (ANM) approaches by shifting from normative inference to participant-specific quantification (Tetreault et al., 2020). While conventional ANM has shown that distributed structural damage converges onto common brain networks, it typically relies on normative connectomes rather than each participant’s own resting-state functional data (Tetreault et al., 2020). This distinction is critical, as functional connectivity exhibits stable individual-specific organization, with particularly high inter-individual variability in heteromodal association cortex (Finn et al., 2015; Mueller et al., 2013). By anchoring each amyloid-positive participant’s seed-based connectivity to the distribution observed in amyloid-negative peers, *π*_*z*_ provide a personalized and statistically grounded measure of deviation from normative functional architecture.

Our primary finding is the emergence of a consistently deviated network despite marked heterogeneity in the anatomical location of individual atrophy seeds. This indicates that, although early structural damage in preclinical AD is spatially diverse, its functional consequences converge onto a more stereotyped system-level organization. This interpretation is consistent with prior work showing that AD-related atrophy follows the architecture of large-scale functional networks across clinical phenotypes (Grothe et al., 2016; Ossenkoppele et al., 2015), as well as with recent evidence demonstrating that preclinical AD atrophy can be localized to a common network using normative atrophy network mapping(Lee et al., 2025). Importantly, our results extend this literature by introducing an individualized perspective, showing that such convergence can be detected directly from each participant’s own functional data, rather than being inferred solely from a normative connectome.

The preferential involvement of the DMN is biologically plausible and consistent with a substantial body of preclinical AD literature (Cha et al., 2026; Guzmán-Vélez et al., 2022; Hampton et al., 2020; Lan et al., 2025; Ossenkoppele et al., 2022). In our data, functional deviations did not distribute randomly across the brain but instead converged onto a coherent large-scale network, predominantly involving transmodal systems, with the DMN showing the strongest contribution (≈ 31% network-referenced overlap and ≈ 30% mask-referenced overlap). This convergence was accompanied by additional involvement of limbic and frontoparietal systems, while primary sensory-motor networks showed only minimal participation. Importantly, these deviations were largely negative (*π*_*z*_ < 0), indicating reduced functional connectivity relative to the A^−^reference, and suggesting a system-level disruption rather than focal alterations.

This preferential engagement of transmodal systems is further supported by network-level enrichment analyses. Considering only effects above the null expectation (E_*n*_ > 1), *π*_*z*_ maps showed maximal enrichment in the DMN (E_*n*_ ≈ 2.0) and secondary enrichment in the limbic network (E_*n*_ ≈ 1.3), reinforcing the idea that functional deviations relative to the normative reference selectively target these systems. Notably, SBC maps, capturing within-group connectivity patterns in A^+^ participants, also showed enrichment in the DMN (E_*n*_ ≈ 1.6) and ventral attention network (E_*n*_ ≈ 1.4), consistent with a distributed reorganization at the functional level. In contrast, W-score maps, reflecting purely morphometric effects, were primarily enriched in the visual network (E_*n*_ ≈ 1.9), with limited contribution from higherorder systems. Together, this pattern highlights a dissociation between structural and functional organization, whereby atrophy remains relatively localized to posterior sensory regions, while its functional impact propagates across large-scale associative networks, particularly the DMN and limbic systems.

This pattern aligns with prior evidence showing that, even in cognitively unimpaired individuals, amyloid burden is associated with abnormal DMN connectivity (Ingala et al., 2021), and in particular in the precuneus (Aponte et al., 2025), and that reduced restingstate integrity within this network predicts subsequent cortical thinning in DMN regions (Hampton et al., 2020). Additional work has suggested that altered interactions between the DMN and control systems mediate the relationship between amyloid pathology and episodic memory performance (Zhukovsky et al., 2023). Within this context, the convergence of individualized functional deviations onto DMN-centered systems supports the view that early AD pathology selectively targets transmodal hubs and their associated networks.

Mechanistically, this interpretation is compatible with emerging models of amyloid-related hyperexcitability, which propose that early increases in neuronal activity —particularly within DMN regions—may drive downstream network instability and vulnerability. This framework is supported by recent work linking DMN hyperexcitability to medial temporal dysfunction and early tau accumulation (Giorgio et al., 2024), as well as broader evidence implicating neuronal hyperexcitability as a key process in AD pathophysiology (Targa Dias Anastacio et al., 2022). In this light, our findings suggest that the observed large-scale functional deviations may reflect the systems-level expression of these early pathological processes, extending beyond the anatomical location of atrophy to impact distributed functional architecture.

Interestingly, *π*_*z*_ maps in A^+^ participants were not significantly associated with continuous clinical variables, including pTau217 levels, Centiloid burden, or PACC performance at time of recruitment. This lack of association suggests that the functional deviations captured by *π*_*z*_ may reflect a network-level state that does not scale linearly with contemporaneous biomarker load or subtle cognitive variation in cognitively unimpaired individuals. This interpretation is consistent with previous work showing that relationships between amyloid pathology, functional connectivity, and cognition in preclinical AD are often complex and non-linear (Ingala et al., 2021; Xu et al., 2024). One possibility is that the transition to amyloid positivity induces a relatively stable reorganization of largescale functional networks, leading to a step-like shift in network dynamics rather than a gradual progression with increasing pathology. Alternatively, functional disruption linked to early structural compromise may reach a plateau during the preclinical phase, limiting the detectability of further variation as amyloid or tau burden increases within cognitively normal cohorts. The absence of associations with PACC may also reflect a temporal dissociation between network-level alterations and the later emergence of measurable cognitive decline, as suggested by longitudinal studies in biomarker-positive but cognitively unimpaired populations (Sperling et al., 2024).

At the same time, the observation that *π*_*z*_ did not relate to standard preclinical cognitive measures contrasts with its robust alignment with canonical cognitive gradients. Specifically, atrophy-related functional deviations were significantly organized along the unimodal–transmodal axis (CogPC1) and the external–internal processing axis (CogPC2), with preferential involvement of transmodal, internally oriented systems. At the level of cognitive concepts, this organization was reflected in significant associations with affective and internally driven processes (e.g., valence, emotion, recognition, retrieval, meaning), alongside negative associations with action- and motor-related domains. This dissociation suggests that *π*_*z*_ captures structured, cognitively meaningful functional variation that could be not well indexed by current gold-standard preclinical cognitive scales. However, there is a considerable number of works that have found associations and predictability between other data domains and cognitive decline. Higher baseline PET centiloids and pTau217 levels were found to be associated with a faster rate of cognitive decline as measured by PACC (Khorsand et al., 2026; Sperling et al., 2024). Another work found that modifiable lifestyle factors such as low education, diabetes, high cholesterol and physical inactivity interact with amyloid status, accelerating PACC decay (Hsu et al., 2026), which aligns with other findings pointing to elevated pulse pressure predicting greater decline in PACC scores (Jung et al., 2025). A multimodal stochastic gradient boosting method including plasma pTau217, baseline PACC, structural MRI features derived from FreeSurfer and tau PET where applicable (Devanarayan et al., 2025) succesfully predicted mean PACC trajectories; and increassed white matter hyperintensities volume in the frontal and parietal lobe were associated with poorer PACC scores (Morales et al., 2024). However, we have not found any work associating functional changes in preclinical stages to PACC performance. In this context, our findings raise the possibility that existing cognitive assessments may lack sensitivity to the types of network-level functional alterations emerging in preclinical AD, and highlight the need for refined or alternative measures that better align with the underlying large-scale functional organization of the brain.

Some limitations and methodological considerations should be acknowledged. The present analyses are primarily cross-sectional, which limits causal interpretation and constrains claims about prognostic value. The clinical relevance of *π*_*z*_ will need to be determined in longitudinal analyses testing whether baseline deviation predicts subsequent biomarker progression or cognitive decline. A relevant methodological consideration concerns the potential first-degree bias described recently in atrophy network dysconnectivity studies (Van Den Heuvel et al., 2026), whereby highly heterogeneous atrophy seeds can give rise to an apparent common network dominated by high-degree hub regions. In this context, it is important to note that *π*_*z*_ maps intrinsically control for this effect, as they explicitly normalize each participant’s connectivity profile against an A^−^ reference, effectively removing the first-order, degree-driven component. As a result, *π*_*z*_ emphasizes networkspecific deviations beyond this generic hub structure. In contrast, raw SBC maps may still reflect this first-degree bias unless explicitly contrasted across groups. Therefore, the *π*_*z*_ framework provides a more specific characterization of functional alterations, isolating deviations that are not trivially explained by network degree or seed heterogeneity.

In addition, *π*_*z*_ exhibited robust alignment with low-dimensional cognitive gradients derived from the Cognitive Atlas and Neurosynth, a structured framework that formalizes relationships between mental processes and experimental tasks. While these gradients (CogPC1 and CogPC2) do not capture the full variability of all cognitive concepts, they provide a principled summary of large-scale cognitive organization. Their association with *π*_*z*_ suggests that functional deviations are cognitively meaningful but not well indexed by current gold-standard preclinical measures, highlighting a potential limitation in the sensitivity of existing cognitive assessments to early network-level alterations.

The temporal trajectory of preclinical AD is generally characterized by the early accumulation of pathological proteins, particularly amyloid-*β* and tau detected through PET imaging, followed only later by the emergence of detectable structural atrophy and measurable cognitive decline. Within this sequence, our *π*_*z*_ approach may capture an intermediate stage between molecular protein deposition and overt neurodegeneration. Specifically, by characterizing functional networks connected to individualized atrophy seeds derived from normative connectivity, *π*_*z*_ may reveal how pathological processes begin to perturb large-scale brain networks before atrophy becomes sufficiently advanced to produce clear cognitive impairment.

Importantly, if *π*_*z*_ simply reflected established atrophy-related degeneration, significant associations with cognitive measures such as the PACC would be expected. However, this was not observed, supporting the interpretation that *π*_*z*_ is sensitive to earlier network-level dysfunction occurring between protein accumulation and macroscopic structural damage. In parallel, we observed positive SBC-related deviations involving frontoparietal regions, particularly the dorsolateral prefrontal cortex (DLPFC), a region frequently implicated in compensatory mechanisms in preclinical AD. Although these positive effects did not survive correction for multiple comparisons (Figure 2), they may nonetheless suggest early compensatory recruitment of higher-order cognitive networks. Future work should determine whether *π*_*z*_ improves risk stratification or predicts the transition from preclinical pathology to measurable cognitive decline.

In conclusion, our results show that the functional impact of structural atrophy in preclinical AD is not spatially random, but instead converges onto a consistently deviated network relative to a healthy reference. By integrating participant-specific atrophy seeds with individual functional connectivity and normative standardization, the *π*_*z*_ framework enables the quantification of individualized network vulnerability beyond conventional group-level approaches. Although these deviations do not scale linearly with contemporaneous amyloid or tau biomarkers, our results suggest that they capture a stable and biologically meaningful signature of early network disruption. As such, *π*_*z*_ provides a promising framework to bridge structural damage and large-scale functional organization, offering new avenues for the characterization of preclinical disease states and the development of network-level biomarkers.

## 4.1. Author Contributions

**Iñigo Tellaetxe-Elorriaga**: Conceptualization, Methodology, Data Curation, Formal Analysis, Visualization, Writing (original draft, review & editing). **Antonio Jiménez-Marín**: Conceptualization, Methodology, Writing (review & editing). **Ibai Díez**: Methodology, Data Curation, Writing (original draft, review & editing) **Jesús Cortés**: Methodology, Writing (original draft, review & editing). **Asier Erramuzpe**: Methodology, Writing (original draft, review & editing).

## 4.2. Acknowledgements

JMC acknowledges financial support from the Spanish Ministry of Health (PI22/01118) and Basque Ministry of Health (2023111002 & 2022111031). JMC and AE are both funded by the Spanish Ministry of Science (grant PID2023-148008OB-I00). ID was supported by the Spanish Ministry of Science with the grant PID2023-150633OA-I00. AE and ID are both funded by the Spanish Ministry of Science and Innovation grants RYC2021-032390-I and RYC2022-035429-I, respectively. JMC, AE, and ID are funded by Ikerbasque: The Basque Foundation for Science.

## 4.4. Data Availability

The A4/LEARN data are publicly available online at the Image and Data Archive (https://ida.loni.usc.edu).

## 4.5. Code Availability

The code corresponding to all the analyses in this study can be found at https://github.com/compneurobilbao/deviation_maps_a4learn and https://github.com/compneurobilbao/compneuro_tools.

